# CTGF is a Modifiable Mediator of Flow-dependent Atherosclerosis under Peripheral Artery Disease Conditions

**DOI:** 10.1101/2023.04.25.23289120

**Authors:** Feifei Li, Sandeep Kumar, Anastassia Pokutta-Paskaleva, Victor Omojola, Dong-won Kang, Chanwoo Kim, Julia Raykin, Carson Hoffmann, Maiko Teichmann, Jing Ma, Hiromi Yanigasawa, Andrew Leask, Lucas Timmins, Xiangqin Cui, Roy Sutliff, Rudy L. Gleason, Hanjoong Jo, Luke Brewster

## Abstract

**Background:** Peripheral arterial disease (PAD) is the 3^rd^ leading type of atherosclerotic disease (ASD) morbidity. Arterial stiffness is intimately connected to the onset and progression of peripheral artery disease (PAD). The role of arterial stiffening on flow-mediated atherosclerotic plaque formation is not well understood. The objective of this study is to discover endothelial cell (EC) pathways under PAD conditions and test the modifiability of these pathways on ASD.

**Methods:** PAD conditions in mice were conferred by partial carotid ligation to induce disturbed Flow (D-flow) in pre-stiffened Fibulin-5 knockout (KO) mice that lack normal elastin function. EC pathways, including Connective tissue growth factor (CTGF/CCN) were quantified by gene analysis and histology. Atherogenic mice had PCSK9 infection + high fat diet. CTGF was inhibited in an EC-specific knockout (ECKO^CTGF^) and with a CTGF antibody (FG-3149). Human vascular tissue was used to validate PAD biomechanics and CTGF upregulation.

**Results:** Biomechanical testing demonstrated that d-flow KO arteries mimic biomechanics of PAD arteries. RNA microarray, qPCR, and immunohistochemistry identified EC plasticity in these arteries compared to WT and KO under stable flow. Under atherogenic conditions, KO arteries demonstrated vulnerable plaques not seen in WT animals. CTGF expression was increased by d-flow, in KO arteries, in aged (18 months) WT arteries, and vascular tissue under d-flow. CTGF inhibition by ECKO^CTGF^ favorably improved plaque characteristics in male but not female animals. FG-3149 treatment of ECWT^CTGF^ male animals delivered similar benefits to plaque characteristics and arterial compliance.

**Conclusion:** ECs in PAD arteries exist under a complex hemodynamic environment that integrates stiffness and d-flow into an atherogenic and inflammatory environment. Stiffness + d-flow stimulates precocious onset of EC plasticity and a vulnerable plaque phenotype. CTGF is a matricellular protein that can tune fibro-inflammatory pathways. CTGF is a prominent mediator of D-flow-mediated arterial remodeling and focal atherosclerotic plaque remodeling. Inhibition of CTGF improves plaque phenotype and arterial compliance. CTGF-associated pathways hold promise as therapeutic targets for PAD patients.

## Clinical Perspective

What is new?

- This study rigorously characterizes a biomechanical model of peripheral arterial disease (PAD) for endothelial cells (ECs) and establishes the unique molecular pathways for ECs under PAD (stiffness + disturbed flow) conditions.
- This study identifies that changes to EC plasticity occur rapidly under PAD conditions and leads to vulnerable plaque profile under atherogenic conditions.
- This study identifies CTGF upregulation in stiffness (in a knockout animal and with aging), that this is increased by disturbed flow, and in both PAD arteries and carotid endarterectomy samples exposed to disturbed flow.
- This study identifies that CTGF inhibition can favorably modulate flow-mediated arterial stiffening and atherosclerotic plaque composition.

What are clinical implications?

- EC plasticity may play an important role in PAD progression and atherosclerotic plaque formation
- Early interventions with CTGF targeted therapies may improve arterial health and atherosclerotic plaque profiles in PAD

## Introduction

Peripheral arterial disease (PAD) is a significant age-related medical condition that is increasing in incidence and impacts 12-20% of the population over 65.^1^ PAD is the 3^rd^ leading type of atherosclerotic disease (ASD) morbidity^2^ and increases the risk of cardiovascular events and mortality 2-3 fold.^3, 4^ ASD is the most common feature of PAD and current medications that target conventional ASD risk factors like hypercholesterolemia, diabetes, hypertension have clear benefit for patients with symptomatic PAD.^5^ However, targeting non-conventional ASD risk factors are the next frontier in clinical management of patients.^2^ As such, the discovery of druggable targets of non-traditional pro-atherogenic pathways is an important endeavor.

Targeting inflammatory pathways may favorably prevent pathologic endothelial cell (EC) plasticity. Both low and oscillatory wall shear stress, termed disturbed flow (d-flow) and arterial stiffening independently promote EC fibrotic and inflammatory pathways.^6, 7^ Recent findings from our group revealed that d-flow patterns in regions of atherosclerotic plaque dramatically alter ECs into proinflammatory, endothelial to mesenchymal (EndMT) and immune cell (End-IT) transition phenotypes in a process defined as endothelial reprogramming (EndRep) that contributes to ASD.^8^ Thus, PAD biomechanics (stiff + d-flow) may stimulate EC inflammation in a synergistic manner that promotes a pro-inflammatory vicious cycle, leading to the distinct differences in atherosclerotic plaque characteristics in PAD compared to coronary artery disease.^9–13^ Thus, mechanically-motivated PAD models may help discover and test PAD-centric molecular pathways that are a currently unmet need.^14, 15^

Under healthy conditions, infra-inguinal arteries are naturally stiffer than elastic arteries.^16^ Stiffened arterial biomechanics are pathognomonic for PAD, and arterial stiffness contributes to both PAD and ASD more broadly.^17, 18^ This stiffening appears to be an important contributor to PAD and PAD symptoms.^19^ As such, baseline stiffened arterial mechanics may influence the onset and progression of PAD in these arteries.^20–22^ The fibulin-5 (Fbln5) knockout mouse (KO) is an ideal murine model of arterial stiffening.^23^ This KO has a deficiency in elastin-based arterial wall mechanics,^24–26^ and in non-diseased states, KO carotid arteries are circumferentially and axially stiffer than arteries from wild-type (WT) mice.^27^

We have found that d-flow induces stiffens otherwise healthy arteries through flow-sensitive matricellular proteins that modulate fibrosis and fibro-inflammation, including thrombospondin-1 (TSP-1).^7^ The fibro-inflammatory molecular pathways particular to PAD biomechanics, and by which EC plasticity is induced in PAD, are not well understood, but likely very important to the development of novel andAD-centric therapies. We hypothesize that it is the combination of arterial stiffening and d-flow that promotes the most pernicious ASD, and that certain fibrotic and/or fibro-inflammatory pathways mediate the pathologic EC plasticity induced under PAD conditions. To test this hypothesis, the objectives of this study were to: 1) develop a murine model of PAD mechanics (stiff + d-flow) and determine the impact of these PAD mechanics on EC pathways and plasticity; 2) quantify differences in flow-mediated atherosclerotic plaque in stiffened KO versus wild type (WT) arteries in atherogenic mice; 3) identify prominent fibrotic mediator (CTGF/CCN2) in ECs from KO and aged mice under d-flow conditions, and in vascular tissue exposed to d-flow (PAD arteries and carotid endarterectomy samples); 4) investigate modifiability of CTGF signaling on arterial stiffness and atherosclerotic plaque in an EC-specific knockout (ECKO^CTGF^) compared to WT control (ECWT^CTGF^) male and female mice; 5) Finally, to test the translational potential of a CTGF antibody (FG-3149) in WT (ECWT^CTGF^) mice.

## Methods

### 1) Animal Handling

All mouse studies performed here were approved by the Institutional Animal Care and Use Committee (IACUC) at Emory University and under the established guidelines and regulations consistent with federal assurance. Fibulin-5 knockout (KO) [C57Bl/6 x 129S2/SvPas] mice were generated from a heterozygous breeding pair originally obtained from Dr. Hiromi Yanagisawa through MTA with UT-Southwestern.^26^ Wild-type littermates were used as control. Constitutive endothelial cell (EC)-specific CTGF knockout (CTGF^fl/fl^; Cdh5 Cre^+/-^) mice were generated by breeding CTGF^fl/fl^ mice (generously provided by Dr. Andrew Leask) with transgenic Cdh5-cre^7Mlia^ mice (Jackson Lab). The CTGF^fl^ allele was converted to null allele by EC-specific Cdh5 Cre-mediated excision. ECWT^CTGF^ (CTGF^fl/fl^; Cdh5 Cre^-/-^) littermates were used as control group. The expression of Cre recombinase in endothelial cells was validated by breeding the Cre mice with Ai14 reporter mice (B6.Cg-Gt (ROSA)26Sortm14(CAG-tdTomato)Hze/J, Jackson Lab). ECKO^CTGF^ (CTGF^fl/fl^; Cdh5 Cre^+/-^) mice showed no difference in an initial clinical and biological analysis at baseline, including body weight and survival. Supplemental Figure 1 provides an overview of the animal experimental methodology and time points for arterial remodeling studies.

### 2) Partial Carotid Ligation Model of D-flow

In the partial carotid ligation model (PCL), three of the four caudal branches of the left carotid arteries (LCAs) were ligated as published,^28^ resulting in a d-flow in LCAs.

Right carotid arteries (RCAs) were not manipulated and had stable flow (s-flow). PCL was performed on all mice groups as listed in Supplemental Figure 1. The in vivo hemodynamic environment in the carotid arteries was characterized at 2 and 4 weeks after PCL using ultrasonography. Blood pressure (BP) was assessed in all murine groups using the CODA noninvasive BP system (a tail-cuff method, Kent Scientific Corporation).

### 3) Biomechanical Testing of Arterial Stiffness

To compare the biomechanical properties, the carotid arteries were harvested from mice quickly after CO_2_ asphyxiation, perfused with saline, isolated, excised, and frozen, prior to being mounted on an ex vivo bioreactor/biomechanical testing device for cylindrical biaxial biomechanical testing as published.^27^

### 4) Endothelial Cell Function in vivo

Age and sex-matched KO and WT carotid arteries were isometrically mounted on wires, placed in an organ chamber containing Krebs-Henseleit buffer (pH is 7.4 when bubbled with 95% O2, 5% CO2 at 37 °C) and connected to a Harvard apparatus differential capacitor force transducer. For each carotid, tension was adjusted to 7.5 mN and then returned to a resting tension 5 mN that yielded maximal contractile response to 50 mM potassium chloride. Data are recorded using PowerLab digital acquisition and analyzed using Chart Software. Results are expressed as mean±SEM. Following precontraction with 2-5 μM phenylephrine, a concentration which yields 80-90% maximum contraction, relaxation responses were examined in response to methacholine (1 nM to 100 M), and the NO donor, sodium nitroprusside (SNP; 1 nM to 100 M). Relaxations were calculated as a percent of the pre-constricted tone induced by phenylephrine. Statistical analyses were performed by two-way ANOVA with Tukey’s post-test repeated measures.

### 5) EC-enriched RNA Isolation from Carotid Arteries

Total RNA from intima (EC-enriched) was collected from KO and WT LCAs and RCAs 24 hours post partial carotid ligation as published.^7^

### 6) Microarray Procedure, Data Analysis, and Bioinformatics

EC enriched RNA from 9 LCAs and 9 RCAs in KO animals were pooled for microarray analysis (n=3 using total of 9 mice per group). RNA sample quality was confirmed, and then samples were amplified and analyzed as a microarray using the MouseWG-6 v2 expression BeadChip array by the Emory Winship Cancer Genomics Core. The normalized microarray data was analyzed statistically, and the statistically significant genes that were differentially expressed genes (LCA vs. RCA) were screened for greater than 1.5 folds increase. The pathway enrichment analysis was performed by mapping these differentially expressed genes to GeneGO’s MetaCore™. This program functions as an integrated software suite for functional analysis of signaling pathways. GeneGO pathways were enriched by the 0.01 gene list derived from the EC-enriched microarray data. The meta-data was uploaded to the GEO Repository (GSE222583).

### 7) En face staining for EC Plasticity

Three days after LCA partial ligation, mice were euthanized by CO_2_ inhalation and perfused with saline containing heparin, followed by a second perfusion with 10% formalin. LCA and RCA were carefully cleaned and dissected free of surrounding tissues and fats. LCA and RCA were en face stained with VE-cadherin antibody (BD Biosciences) and α-SMA antibody (abcam). LCA and RCA were counterstained using DAPI (Sigma) and mounted on glass slides using fluorescence mounting medium (Dako). En face images were collected with a Zeiss LSM 510 META confocal microscope.

### 8) Murine Model of Atherosclerosis

Atherosclerosis in left carotid arteries was induced in all murine groups using male and female mice aged 12-20 weeks old as published.^29^

### 9) Histological and Immunohistochemical Analysis

Oil red O (ORO), H&E, Masson’s trichrome staining, and elastin autofluorescence were conducted on sections of common carotid arteries. Lipids were detected with ORO (Sigma) staining following the standard protocol as described.^28^ Masson’s trichrome staining was used for the assessment of collagen composition of the atherosclerotic plaque. Elastin architecture was visualized by autofluorescence.

Immunohistochemical (IHC) staining of CD68 (BioRad) was applied to quantify macrophage positive area. The quantitative assessments of the arterial wall and necrotic area were blinded and quantified using Image J program. Each value was based on 3 replicates. Unpaired, two-tailed t-test was used to compare mean values of comparison d-flow carotid arteries with statistical significance at P<0.05.

### 10) Human Tissue Validation

Human artery samples from PAD and carotid endarterectomy (CEA) patients (Supplementary Tables 1 and 2) were collected from operative specimens after informed consent under an IRB approved protocol (Emory University approved IRB protocol numbers: 51432 and 70813). Arterial specimens were segregated prior to testing into d-flow conditions if there was not inline flow to the artery as validated and published prior^7^. In contrast, s-flow was defined in the same way as murine RCAs and attributed to arteries with inline flow to them. All d-flow arteries were in patients with proximal inflow occlusion and distal reconstitution through collateral pathways.

All s-flow arteries had inline flow without obstructive PAD.

### 11) CTGF Inhibition on Murine Atherosclerosis Model

The effects of CTGF inhibition on atherosclerosis development were examined through EC-specific CTGF knockout and CTGF monoclonal antibody (FG-3149) treatment in carotid atherosclerosis model. FG-3149 was generously donated to the Brewster laboratory by FibroGen Inc. (San Francisco, CA). Male ECWT^CTGF^ mice under atherosclerosis model were assigned to 2 antibody treatment groups: sham antibody (IgG; n=6) and FG-3149 (n=6). Antibodies were administered by intraperitoneal injection twice weekly at 30 mg/kg since day 1 after PCL for one month.

### 12) Statistics

Where not already stated, unpaired t-test were performed with GraphPad Prism (GraphPad Software). Parameters such as sample size, the number of replicates, the number of independent experiments, measures of center, dispersion, and precision (mean ± SD), and statistical significance are reported in figures and figure legends. Statistical significance was set at P<0.05.

## Results

### KO carotid arteries mechanics

To compare KO and WT carotid arteries to PAD arteries, we used the *ϕ_E_* (d-flow/s-flow ratio) of respective murine carotid artery elastic moduli and compared this to that of PAD arteries/aged healthy artery elastic moduli. Here, we found that KO arteries closely aligned with an approximately 3 fold stiffer ratio over physiologic mean pressures that was similar to that characterized in PAD arteries (Figure 1A). To identify the baseline stiffness of KO carotid arteries to that of aged control carotid arteries, we compared the compliance curves of unmanipulated KO arteries to 80-week old (∼18 months old) aged WT control mice arteries. Here we found that the compliance of unmanipulated (stable or s-flow) KO arteries were overall similar to aged WT arteries but even stiffer than aged control arteries from 60-80 mmHg (Figure 1B). To compare the impact of d-flow on KO carotid artery stiffening, we compared pressure-diameter and compliance curves of KO left carotid arteries (LCAs) exposed to d-flow to unmanipulated right carotid arteries (RCAs), which have stable flow (s-flow). After d-flow, KO LCAs underwent profound stiffening in the range of physiologic mean pressures, which is outlined by the box (Figure 1C, D). There were no differences in arterial stiffening in KO female versus male carotid arteries, regardless of flow conditions (Supplementary Figure 2). KO arteries had lower in vivo axial stretches (axial stiffening) with KO d-flow arteries the least compliant (Aged WT>KO RCA>KO LCA; Figure 1E). However, both KO RCAs and LCAs have robust residual stresses that are not further increased under d-flow (Figure 1F). PAD arteries have EC dysfunction. To test EC dysfunction in KO and WT arteries, EC-dependent and independent relaxation of KO and WT carotid arteries was performed. KO (but not WT) carotid arteries exhibited a basal EC dysfunction, indicated by impaired endothelial dependent vasodilation in arterial ring testing (Figure 1G) that was ameliorated by exogenous sodium nitroprusside (Figure 1H).

**Figure 1:**
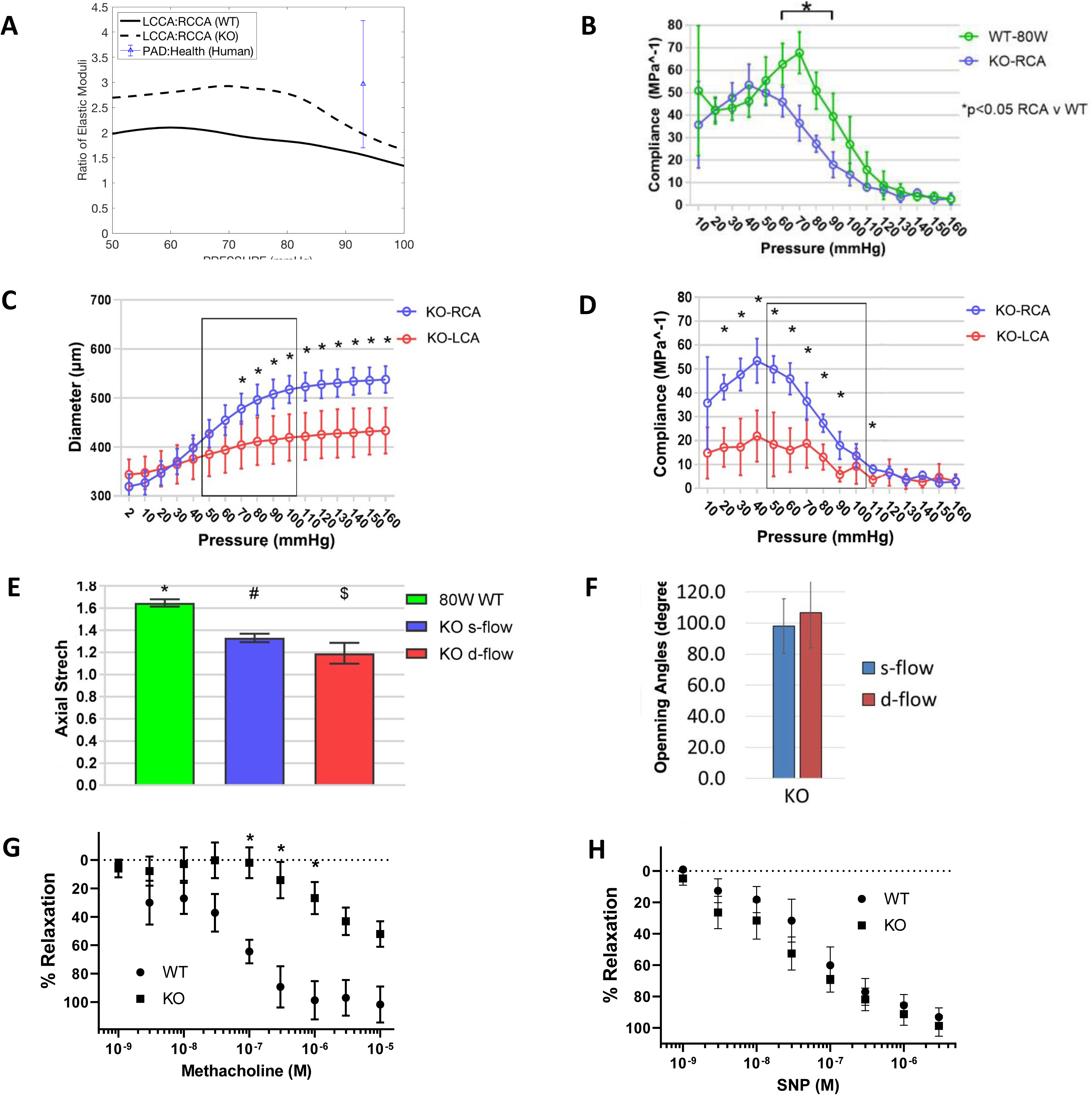
Fibulin-5 Knockout (KO) Mimics the Hemodynamics and EC Dysfunction of PAD. **A**. In order to compare relative biomechanical changes between wildtype (black line) and KO animal arteries (dotted line), an elastic modulus ratio was calculated from the left common carotid arteries (LCA exposed to d-flow) and the right common carotid arteries (RCA exposed to s-flow) over physiologic mean arterial pressures. We then calculated an elastic modulus ratio for PAD arteries compared to healthy arteries. Mean average ratio and standard deviation is superimposed as a triangle and blue line to the right of the curves. The KO artery ratio over physiologic mean pressures is greater than that of WT arteries and very similar to that of PAD arteries (blue triangle). **B.** To compare basal stiffness of KO carotid arteries to that of aged animals (80 weeks; ∼18 months), we compared s-flow KO RCA to that of aged mice. While most of the pressure ranges had similar stiffness, the KO RCA was stiffer than aged arteries from 60-80 mmHg. (KO n= 6-7; 4 males/3 females) **C/D**. To quantify flow-mediated stiffening in KO arteries, pressure-diameter and compliance curves were generated from biaxial testing to demonstrate that the d-flow KO carotid arteries were circumferentially stiffer than the contralateral s-flow RCAs. **E.** To test longitudinal stiffening, aged arteries were compared to KO under s-flow and d-flow. Here there was sequential stiffening with d-flow KO least compliant. Symbols indicate statistical significance. **F.** Residual stress is a measurement of pathologic forces in the arterial wall. Opening angles in both KO RCA and LCA were exuberant but similar. **G/H.** To compare baseline endothelial cell dysfunction, endothelial cell dependent (G, N=6) and independent (H, N=6) relaxation was quantified via arterial ring testing. WT: 2 male, 4 female; KO: 3 male, 3 female. (**B-D:** Unpaired multiple t-tests with Holm-Sidak method; **E:** One-way ANOVA with Tukey’s post hoc test; **G/H:** Two-Way ANOVA with Tukey’s post hoc test (*p<0.05 compared to WT).

### Validating d-flow environment/EC response to d-flow in KO carotid arteries

Next, the fidelity of the d-flow environment over the experimental time course was verified using duplex ultrasound (2/4 weeks). The RCA demonstrates s-flow with laminar WSS, whereas the LCA demonstrates low and oscillatory wall shear stress (Figure 2A-C). To identify early EC-enriched RNA pathways in KO arteries, we examined intimal RNA after 24 hours of d-flow. ECs on the d-flow, KO LCAs had significant downward expression of PECAM-1 and similar α-SMA levels (Figure 2D). By 72 hours of d-flow, en face staining of the intima demonstrates increased expression of α-SMA in the KO LCAs compared to d-flow WT LCAs (Figure 2H, F), as well as compared to both KO and WT RCAs, which were exposed to s-flow (Figure 2G, E). This induction of EC plasticity towards EndMT in KO + d-flow was verified in microarray pathway analysis and compared to that of d-flow, WT arteries (Figure 2I, J). Here the top 5 identified differentially expressed clusters in KO carotid arteries included cytoskeleton remodeling signaling pathways (including TGF and Wnt) along with the EndMT pathways. Venn diagram of microarray data comparing flow mediated differences within the KO and WT arteries identified only 24 overlapping genes, and 138 or 398 gene differences between the groups. This identifies disparate genetic fingerprints between the KO and WT arteries under flow (Supplementary Figure 3A, B; Supplementary Data File).

**Figure 2:**
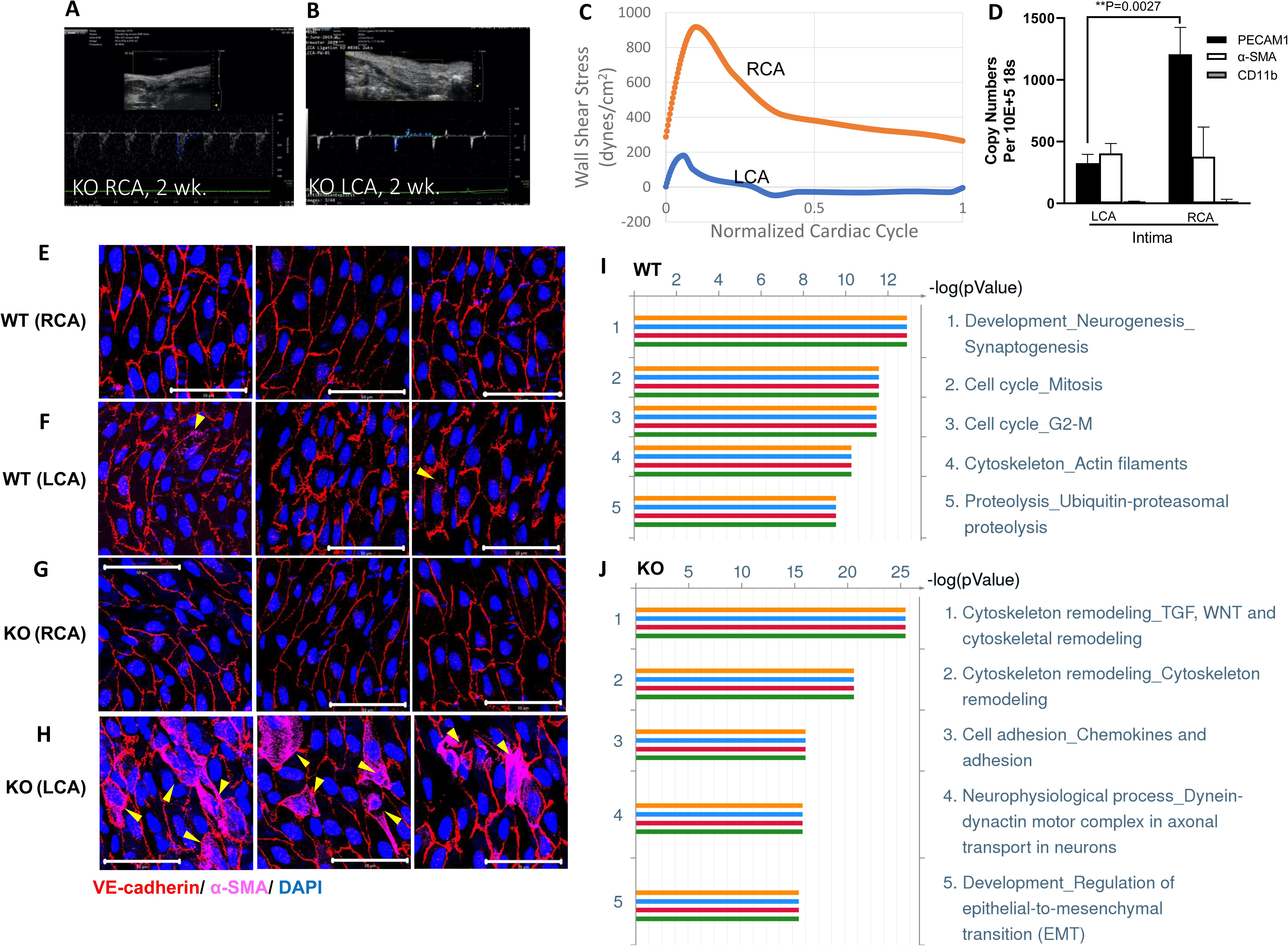
EC Plasticity in KO Carotid Arteries Under D-flow. **A/B.** Representative duplex ultrasonography images of KO RCA and LCA two weeks after PCL. **C.** LCAs with d-flow manifest low and oscillatory WSS throughout the cardiac cycle (blue curve) compared to RCA (orange curve). **D.** 24 hours after PCL, intimal RNA expression of PECAM-1 was reduced in KO LCA endothelium compared to RCA. No significant change was observed in α-SMA and CD11b expression (**D:** Mean ± SD; unpaired t-test with significance set at P<0.05; n=3). **E-H.** En face immunofluorescence staining of the intima demonstrates robust α-SMA expression in the endothelium of KO LCAs (**H**) after 72 hours under d-flow. Scale bar: 50μm. **I/J.** Cluster pathway analysis identified differentially expressed genes (DEGs) in WT (**I**) and KO (**J**) arteries. Cell cycle and changes in the actin cytoskeleton predominated in the WT animals, while EndMT and TGF-beta/Wnt cytoskeletal remodeling was more prominent in KO consistent with cell stiffening and EC plasticity pathways.

### KO carotid artery plaque under d-flow + atherogenic conditions

Since many of the identified fibro-inflammatory pathways that affect EC plasticity and arterial stiffening may also contribute to ASD plaque remodeling, an atherogenic environment was imposed on KO animals +/- d-flow; WT animals under the same conditions served as normal arterial compliance control animals. Histologic examination of carotid artery plaques was performed for weeks after induction of d-flow. Only LCAs under d-flow developed plaques; this occurred in both WT and KO animals. (Representative histology presented in Figure 3A-E) Here, pre-stiffened KO arteries demonstrated a more aggressive, vulnerable atherosclerotic plaque formation in response to d-flow. Specifically, KO LCAs had significantly more plaque area, lipid deposition in the plaque, percent necrotic area in the plaque, and macrophage content in the plaque (Figure 3F-I). KO arteries also had increased inner and outer diameter (Figure 3L-M), consistent with Glagov’s remodeling,^30^ that correlated with an increase in elastin breaks (Figure 3N). In contrast, WT LCAs demonstrated a more fibrotic plaque phenotype with medial thickening and greater collagen content (Figure 3J, O-P). Atherogenic conditions did unmask an increase MAP in WT (compared to KO) male animals that did not exist under standard, non-atherogenic conditions (Supplementary Figure 4C, 3^rd^ panel). There were no differences in the serum lipid levels between KO and WT animals (Supplementary Figure 4E-L).

**Figure 3:**
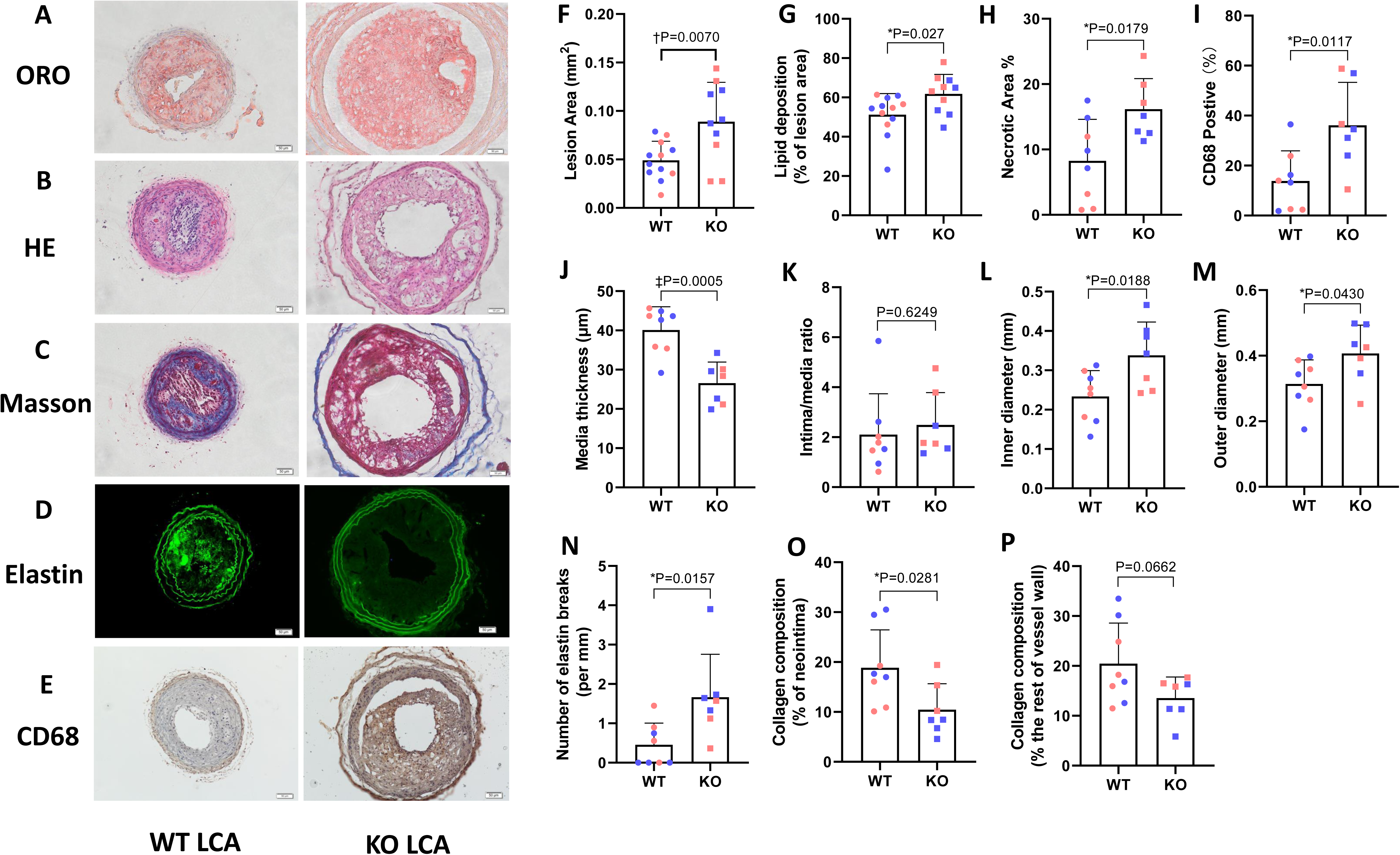
PAD Conditions (Stiff + D-flow) Induce Vulnerable Plaque Phenotype. **A-E.** Representative images of Oil Red O (ORO), H&E, Masson’s trichrome, elastin autofluorescence and IHC staining of macrophages (CD68) of the LCA from WT and KO animals. **F.** KO LCAs showed significant larger atherosclerotic plaque burden. **G.** Lipid deposition within the plaques is higher in KO mice. **H/I.** Plaques in KO mice had increased necrotic area and greater macrophage infiltration**. J/K.** KO arteries had thinner medial layers than WT, but no difference in intima/media ratio. **L/M.** KO LCAs had increased inner and outer diameters of their lumen and arterial walls than WT. **N.** KO LCA also had increased elastin breaks compared to WT LCAs. **O/P.** In contrast, WT LCAs developed more collagen content in the plaque; collagen content in the rest of the arterial wall was similar. WT n=12 (7 males/ 5 females) and KO n=10 (5 males/ 5 females) in ORO quantification; WT n=8 (4 males/4 females) and KO n=7 (4 males/3 females) in the rest quantifications; male and female mice are represented with blue and pink symbols, respectively; data expressed as mean ± SD; unpaired t-test with significance set at P<0.05. Scale: 50 μm.

### CTGF expression in response to aging and d-flow

When examining our WT and KO carotid artery gene expression data under d-flow, we found that CTGF was increased under d-flow and in KO compared to WT arteries. Specifically, and in a stair-step manner, CTGF was upregulated in d-flow WT LCAs compared to s-flow RCAs, and that s-flow KO RCAs had similar CTGF expression to that of d-flow WT LCAs. Further, CTGF expression increases in d-flow KO LCA compared to s-flow KO RCAs (Figure 4A). Since arterial stiffening and PAD is also associated with aging, we queried our young versus aged (18 months) murine s-flow/d-flow WT gene dataset to determine the impact of d-flow and aging on CTGF.^31^ We discovered a similar stair-step relationship for d-flow and aging in the upregulation of CTGF. Here, d-flow upregulates CTGF in young animal carotid arteries compared to s-flow, and older s-flow arteries have similar CTGF expression to that of younger d-flow arteries. Similarly, older d-flow carotid arteries had significantly greater CTGF expression (Figure 4B) than older, s-flow arteries.

**Figure 4:**
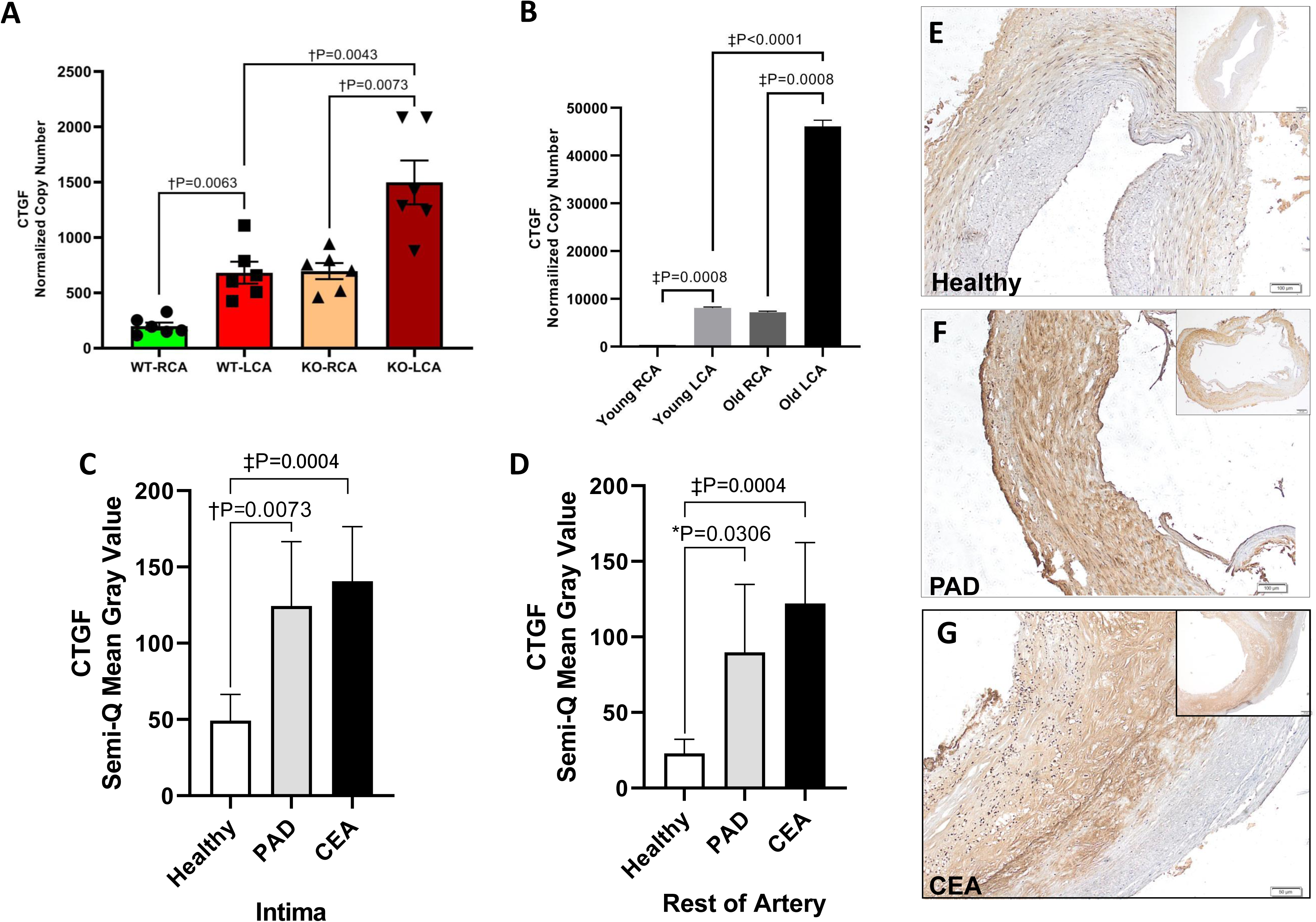
CTGF Expression is Flow-mediated and Upregulated in Stiffened KO and Aged Murine Arteries and Under d-flow in PAD Arteries and Carotid Endarterectomy Specimens. **A.** D-flow increases CTGF expression in LCAs. WT LCA and pre-stiffened, KO RCAs have similar CTGF expression levels, and CTGF is further increased in KO LCA compared to KO RCA. **B.** Similarly, CTGF expression is increased under d-flow in the LCA of young mice over that of s-flow RCAs, and to a similar degree as s-flow RCAs in old mice (80 weeks old). LCAs in older mice under d-flow is significantly increased over that of young d-flow and aged s-flow. **C/D**. The intima and rest of artery of both PAD arteries and CEA plaques under d-flow have increased CTGF deposition over that of aged healthy control arteries, respectively. **E-G.** Representative images of IHC staining of CTGF in healthy aged peripheral arteries, PAD arteries and carotid artery plaque. n=6 for healthy group; n=7 for PAD group; n=16 for CEA; mean ± SD; one-way ANOVA with Tukey’s post hoc test. Scale: 50 μm.

Prior to testing the modifiability of CTGF expression under PAD conditions, we validated the importance of CTGF in human PAD pathology. IHC staining for CTGF was quantified in d-flow PAD arteries and compared to aged infra-inguinal arteries under s-flow (patient information in Supplementary Table 1). To further test the role of d-flow on CTGF expression in vascular patients, we compared CTGF staining in carotid endarterectomy samples (patient information in Supplementary Table 2).

Here too, we found CTGF significantly upregulated both in the intima and remainder of the artery in PAD arteries and carotid endarterectomy samples compared to aged s-flow arteries (Figure 4C-G), establishing CTGF is increased under d-flow in PAD and carotid arteries.

### Targeted CTGF therapies manipulate flow-mediated atherosclerotic plaques

To test the role of EC CTGF in flow-mediated atherosclerotic plaque biology, we created an EC-specific CTGF KO mice strain (ECKO^CTGF^). We imposed atherogenic conditions in the same manner as prior (PCSK9 infection + high fat diet), then created d-flow by PCL. Male ECKO^CTGF^ mice developed less plaque burden and percent macrophage content in plaque under d-flow + atherogenic conditions compared to littermate control animals (Figure 5A, F, I). Female ECKO^CTGF^ mice developed a trend (P=.06) towards decreased necrotic area compared to littermate control animals (Supplemental Figure 5H) but were otherwise similar to control animals. To test the effect of CTGF inhibition with a monoclonal antibody (FG-3149; donated for research testing by Fibrogen, Inc.) as a translational therapeutic, male littermate WT control animals (ECWT^CTGF^) under atherogenic conditions were treated with FG-3149 or control IgG antibody beginning on postoperative day 1 after PCL (induction of d-flow). Again, CTGF inhibition impacted flow-mediated carotid plaque. Specifically, plaque in d-flow LCA of animals treated with CTGF antibody (FG-3149) developed less lipid deposition and percent necrotic area in their plaques compared to IgG (Figure 6A, G-H).

**Figure 5:**
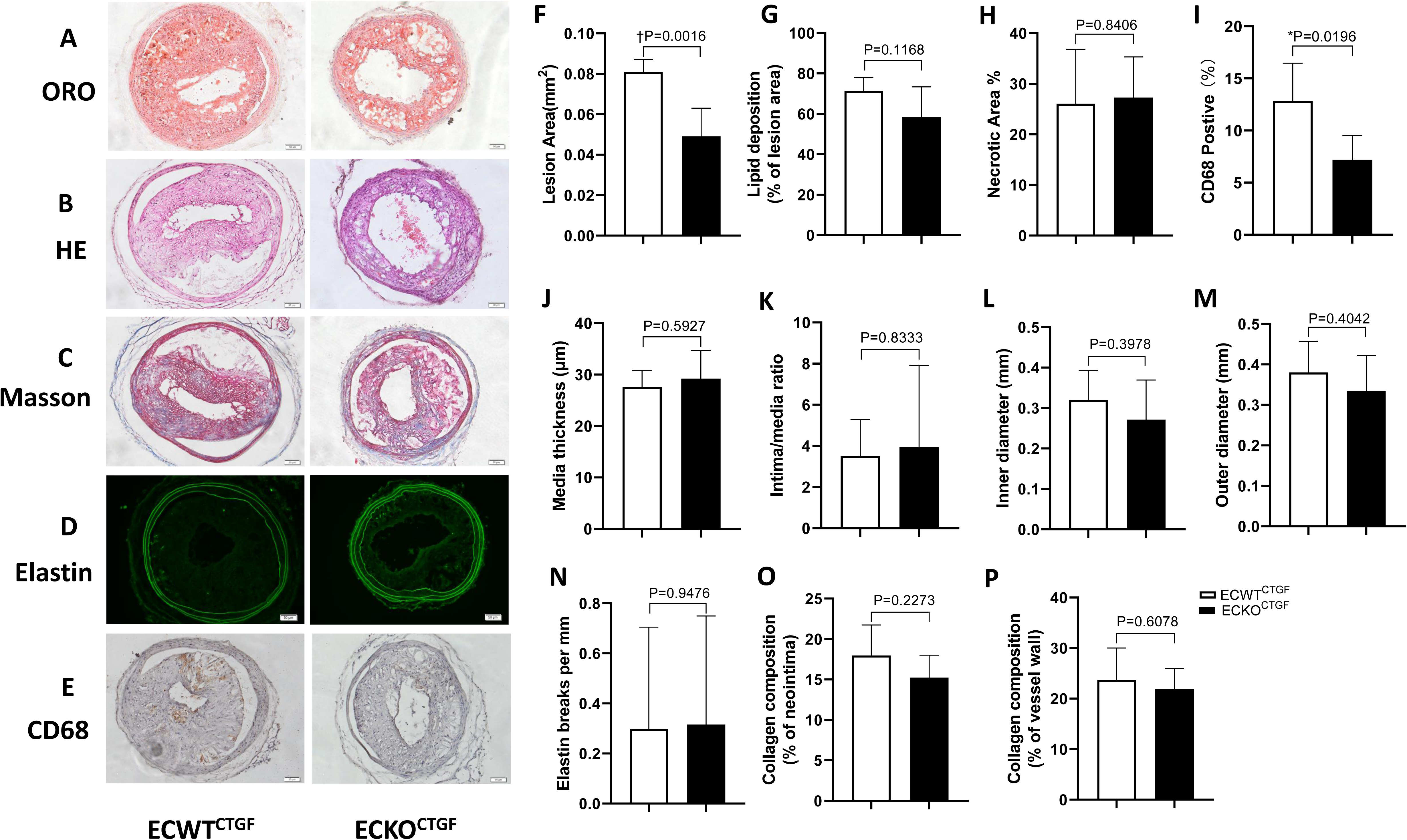
Endothelial Cell CTGF Knockout (ECKO^CTGF^) Decreases Plaque Area and Macrophage Infiltration in Male Mice Carotid Arteries Under D-Flow. **A-E.** Representative images of Oil Red O (ORO), H&E, Masson’s trichrome, elastin autofluorescence and IHC staining of macrophages (CD68) of the LCAs from EC knockout of CTGF (ECKO^CTGF^) and WT (ECWT^CTGF^) animals. **F.** ECKO^CTGF^ mice have less atherosclerotic plaque area than control. **G/H.** Lipid deposition and necrotic area are similar between groups. **I.** ECKO^CTGF^ mice have less macrophage infiltration. **J/K.** ECWT^CTGF^ and ECKO^CTGF^ mice have similar medial thickness and intima/media ratio. **L/M.** The Inner luminal and outer arterial wall diameters of LCAs are similar between groups. **N-P.** There are no differences between groups in elastin breaks or the collagen content of the arterial intima or rest of arterial wall. ECWT^CTGF^ n=5; ECKO^CTGF^ n=5. Data expressed as mean ± SD; unpaired t-test with significance set at P<0.05. Scale: 50 μm.

**Figure 6:**
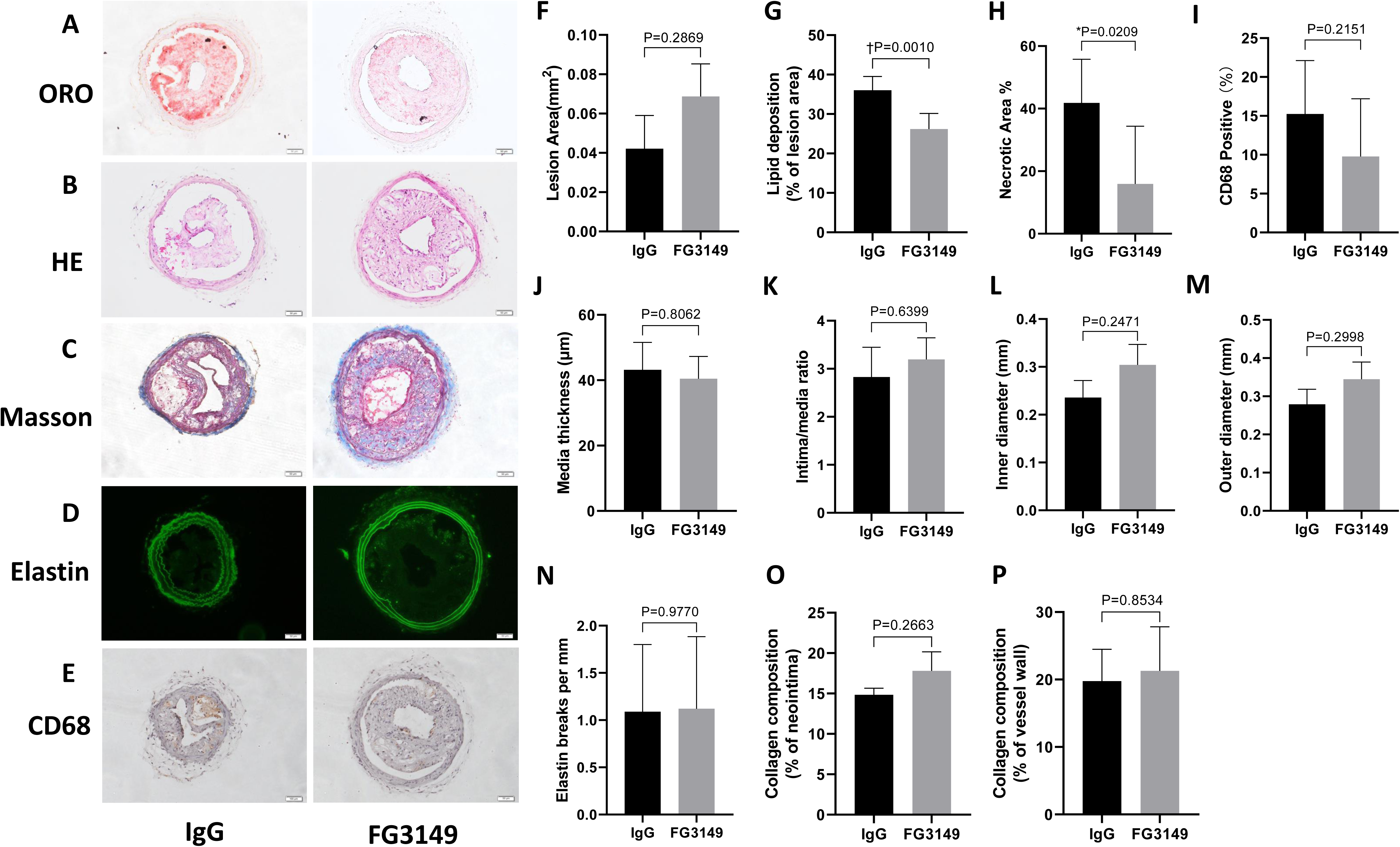
CTGF Antibody (FG-3149) Decreases Lipid Content and Necrotic Area in Male Mice Carotid Arteries Under D-Flow. **A-E.** Representative images of Oil Red O (ORO), H&E, Masson’s trichrome, elastin autofluorescence and IHC staining of macrophages (CD68) of the LCAs. **F.** FG-3149 treatment did not decrease atherosclerotic plaque area change. **G/H.** FG-3149 treatment decreased lipid accumulation and necrotic area. **I.** There was a trend towards decreased macrophage infiltration in the FG3149 treatment group. **J/K.** There were no differences in medial thickness or the intima/media ratio. **L/M.** There were no differences in the Inner luminal or outer arterial wall diameters between groups. **N-P.** There were no differences in elastin breaks or the collagen content within the intima or rest of arterial wall. IgG Sham antibody group n=6; FG-3149 group n=6; mean ± SD; unpaired t-test with significance set at P<0.05. Scale: 50 μm.

### Targeted CTGF therapies improves arterial mechanics during flow-mediated remodeling

To test whether the effect of EC-specific CTGF knockout on artery stiffness, biaxial mechanical tests were applied to carotid arteries from ECWT^CTGF^ and ECKO^CTGF^ after 6 weeks of PCL under non-atherogenic conditions. Differences between LCA/RCA compliance curves are over a narrower pressure range in the ECKO^CTGF^ compared to ECWT^CTGF^ (Figure 7A, B). Similarly, animals treated with CTGF antibody (FG-3149) have similar differences, also over a narrower pressure range compared to IgG control arteries (Figure 7E, F). The axial stiffening of the ECWT^CTGF^ LCA compared to RCA trended towards significance (P=.05), whereas it did not (compliance more preserved) in the ECKO^CTGF^ LCAs (Figure 7C). Axial stiffness was significantly greater in IgG treated animals’ LCA compared to RCA, but not FG-3149 treated arteries, which were protected (Figure 7G). Moreover, under atherogenic conditions, the residual stresses of the ECKO^CTGF^ LCAs are significantly decreased compared to ECWT^CTGF^ LCAs (Figure 7D), supporting a favorable local impact on plaque mechanics.

**Figure 7:**
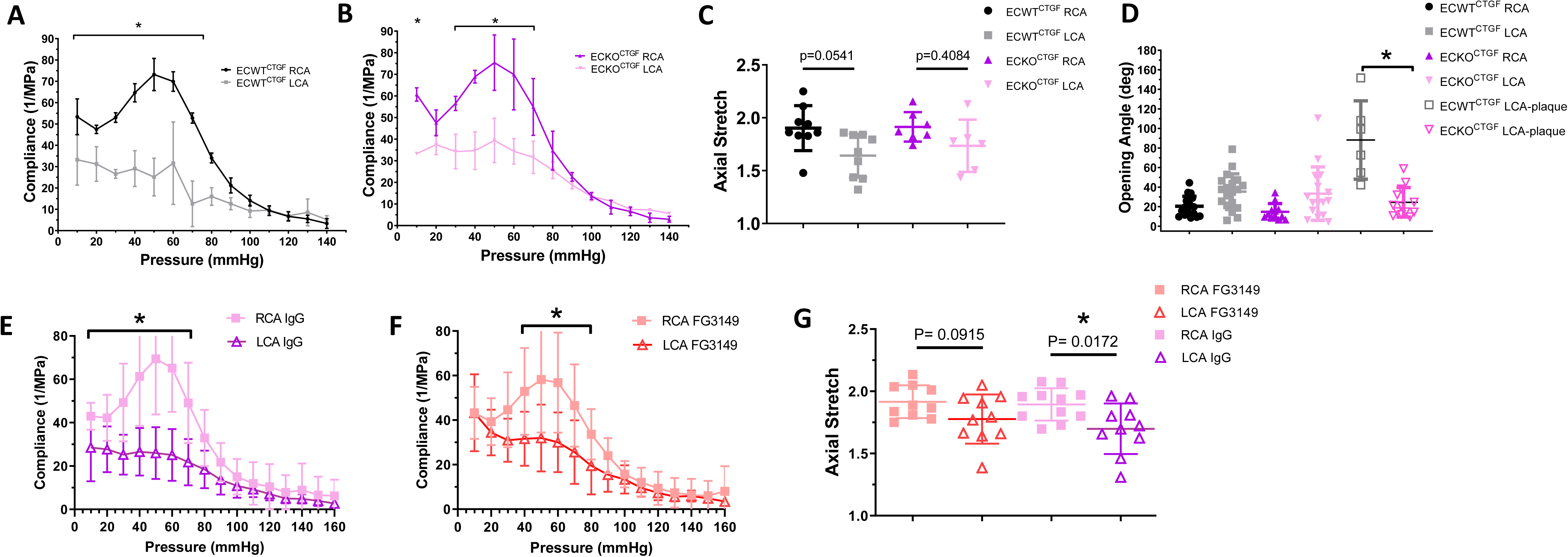
CTGF Inhibition Improves Arterial Compliance During Disturbed Flow Mediated Arterial Remodeling. **A.** Pressure-compliance curve of wild type (ECWT^CTGF^) animals demonstrate decreased arterial compliance in LCAs under 10-80mmHg range (LCA n=4; RCA n=3; all male). Two-way ANOVA w/ Tukey correction. * p<0.05, [p=0.0006 p=0.0102 p<0.0001 p<0.0001 p<0.0001 p<0.0001 p<0.0001 p<0.0001] **B.** Pressure-compliance curves of ECKO^CTGF^ have a narrower range of pressures (10, 30-70 mmHg) that have a significant difference in arterial compliance between the LCA and RCA. (LCA n=3; RCA n=3; all male). Two-way ANOVA w/ Tukey’s posthoc *p<0.05 [p=0.028 p=0.0019 p<0.0001 p=0.0010] **C.** LCA from ECWT^CTGF^ trend toward significant axial stiffening as measured by axial stretch in the LCA versus RCA; axial compliance is preserved in ECKO^CTGF^ LCAs. One-way ANOVA w/ Tukey correction, nonsignificant. n=10, n=9, n=7 n=6 (RCA, LCA, KO-RCA, KO-LCA, respectively); p-values in figure. **D.** There were no differences in the opening angles between RCA and LCA in in the wild type (ECWT^CTGF^: black/grey) or EC CTGF KO (ECKO^CTGF^: purple and pink). Under atherogenic conditions, the wild type ECWT^CTGF^ animal has significantly greater residual stresses compared to ECKO^CTGF^ in the d-flow LCA. ECWT^CTGF^ RCA (n=27); ECWT^CTGF^ LCA (n=24); ECKO^CTGF^ RCA (n=15); ECKO^CTGF^ LCA (n=15); ECWT^CTGF^ LCA-plaque (n=20); ECKO^CTGF^LCA-plaque (n=16). Ordinary one way ANOVA w/ Tukey’s correction, ECWT^CTGF^ LCA-plaque ECKO^CTGF^ LCA-plaque *p<0.0001 **E.** Pressure-compliance curves of carotid arteries from wild type (ECWT^CTGF^) animals treated with sham IgG antibody demonstrate decreased arterial compliance in LCAs under 10-70mmHg range (LCA n=11; RCA n=11; 6 male/5 female). Two-way ANOVA w/ Tukey’s correction *p<0.05 [p=0.0283 p=0.0216 p<0.0001 p<0.0001 p<0.0001 p<0.0001 p<0.0001 p=0.0215] **F.** Pressure-compliance curves of carotid arteries from ECWT^CTGF^ treated with CTGF antibody (FG-3149) have a narrower range of pressures (40-80mmHg) that have a significant difference in arterial compliance between the LCA and RCA (LCA n=10; RCA n=10; 6 male, 4 female). Two-way ANOVA w/ Tukey’s correction *p<0.05 [p=0.0006 p<0.0001 p<0.0001 p=0.0008 p=0.0477] **G.** There is a significant difference in axial stiffening under d-flow (LCA) in the IgG treated animals. This is attenuated and not significant in the CTGF antibody (FG-3149) treated animals. RCA IgG n=11, LCA IgG n=10, RCA FG3149 n=10, LCA FG3149 n=10). Ordinary one-way ANOVA w/ Tukey’s correction. P value in figure.

## Discussion

This study has 5 important findings. First, this work characterized a murine model that mimics the biomechanics of PAD and establishes the powerful effect of biomechanical forces acting on ECs under these PAD conditions (stiffness + d-flow in an athero-inflammatory environment) on EC pathways and plasticity. As such, this work contributes to the emerging collective work linking arterial stiffness and vascular disease.^32, 33^ The addition of d-flow to KO arteries builds on prior work characterizing the stiffened profile of Fbln5 KO arteries previously that was mostly due to elastin dysfunction,^27^ and our prior discovery that d-flow mediated arterial stiffening was from fibrotic pathway upregulation and collagen deposition^7^. Here, we quantify that this combination mimics the mechanical environment of PAD arteries.

EC plasticity is a likely mechanism as we demonstrate rapid EndMT changes in the endothelium of KO mice exposed to d-flow. A single cell RNA (scRNA) sequencing analysis by the Jo lab characterized the effect of d-flow on individual EC plasticity.^8^ Here, d-flow alone, promoted EC plasticity into a wide variety of phenotypes from inflammatory to mesenchymal (i.e., EndMT), immune cell-like, stem/progenitor-like, and hematopoietic phenotypes over ∼14 days. The impact of d-flow under stiffened conditions appears to speed up this process in the murine model (3 days compared to two weeks). This has clinical implications because PAD most commonly presents as a progressive obstructive disease due to focal deposition of atherosclerotic plaque at branch points, where the arteries exhibit d-flow patterns. Thus, conceptually in PAD, this work supports the paradigm that ECs in already stiff PAD arteries and exposed to d-flow, are “tuned” to direct a *unique molecular signature* to d-flow that is more pernicious than that seen in less stiff arteries. This appears to be further tuned by the matricellular protein, CTGF. Since ECs in the endothelium do not act as one, but rather undergo EndRep into non-conventional ECs that can direct focal inflammation and progression ASD. How EC plasticity and matricellular proteins interact, and how can they be harnessed to develop next generation PAD therapies is a promising avenue for future discovery.

Secondly, the combination of stiffness + d-flow leads to this particularly aggressive atherosclerotic plaque remodeling as evidenced by plaque area, plaque content, and elastin breaks. Interventions that limit stiffness (targeted CTGF therapies) or promote s-flow over d-flow (revascularization procedures) may reverse the velocity of atherosclerotic remodeling and maybe outcome of this process, but his remains to be definitively proven. It is highly likely that the effect of PAD conditions (stiff + d-flow) on EC plasticity will be an important surrogate for future therapeutic testing due to the known causative role for EndMT in atherosclerotic plaque formation.^34^ There are also several studies supporting an important role for matrix stiffness on pathologic behavior of other cell types that provides broader support of this concept. For example, In myofibroblasts, matrix stiffness promotes the activation of RhoA,^35^ which is the upstream regulator of transcription factor Twist1,^36^ Snail1 and Slug.^37^ Also, YAP/Taz is recognized as the key regulator in matrix stiffness induced EndMT through collaboration with Snail1/Slug in mesenchymal stem cells. ^38^ Interestingly, YAP/Taz is both upstream and downstream from CTGF,^39^ making CTGF particularly exciting as a therapeutic target. Similar early gene regulation is seen in our EC-enriched gene analysis. (Supplementary Data File)

Thirdly, we identified CTGF as a prominent target as a fibrotic mediator of stiffened arterial remodeling under d-flow conditions, and in both KO and aged animals. We validated CTGF was upregulated in ECs from KO and aged mice under d-flow conditions, and in vascular tissue exposed to d-flow (PAD arteries and carotid endarterectomy samples). CTGF is a multifunctional matricellular protein that is composed of 4 modules, i.e., IGFBP (insulin-like growth fac-tor binding protein)-like, VWC (von Willebrand factor type C), TSP1 (thrombospondin 1 type 1) repeat, and CT (C-terminal cystine knot) module. All 4 of these modules comprising CTGF are highly interactive with a variety of other molecules such as growth factor binding, integrin recognition, and interaction(s) with heparin and proteoglycans. CTGF has an important role in cell adhesion, migration and proliferation, which is correlated to the initiation and progression of atherosclerotic lesions in humans and in animal models.^40^ Although CTGF is highly expressed in complicated compared with fibrous plaques,^41^ no clear mechanisms have been attributed to CTGF in atherosclerotic plaque. Our work here supports this may be due in part to favorable modulation of arterial compliance.

Fourth, in order to test the modifiability of CTGF signaling on arterial stiffness and atherosclerotic plaque composition, we induced d-flow in an EC-specific knockout (ECKO^CTGF^) compared to WT control (ECWT^CTGF^) male and female mice under standard and atherogenic conditions. Here we identified a critical and modifiable role for CTGF in both arterial stiffening and atherosclerotic plaque characteristics. To our surprise, the favorable plaque findings appeared to be limited to male animals. Here, we discovered that CTGF ECs specific knockout (ECKO^CTGF^) have decreased atherosclerotic plaque size in male mice compared with littermates. This was not seen in female mice. Due to these findings, we limited our testing of CTGF mAb to male littermate control mice (ECWT^CTGF^) tested against IgG treated control animals. Here we found FG-3149 decreased lipid content and necrotic area in plaques. Sex-based differences in PAD is an important area of research, and it is possible that matricellular proteins, like CTGF, could be differentially regulated in this manner. Sex differences may impact the benefits of CTGF inhibition on atherosclerotic plaque biology.

While the burden of PAD with increasing age is shared by both sexes,^42^ clinically, PAD in women is delayed with age proportionately to the delayed onset of arterial stiffness and EC dysfunction, ∼10 years after men;^42^ delays in stiffening are also seen in murine models.^43^ Arterial stiffness is of particular interest to sex-based differences because not only is it a risk factor for CV disease and mortality,^44^ but the association between arterial stiffness and mortality is ∼2 fold greater in women.^45^ In this manuscript, we focused on the EC response to PAD mechanics, but further sex-based investigation of matricellular pathways under PAD conditions is warranted. Finally, we tested the translational potential of a CTGF antibody (FG-3149) in WT (ECWT^CTGF^) male mice. Here too, we found favorable modulation of atherosclerotic plaque with our antibody therapy. There is likely an important interaction between CTGF modulation of fibro-inflammatory pathways induced by d-flow. We know that d-flow induces proinflammatory gene expression and induces atherosclerosis development.^46^ Here, we uniquely have identified that CTGF increases in d-flow, in KO arteries, and in older arteries (that have similar mechanics to KO arteries under s-flow), and validated that CTGF is increased under d-flow conditions in PAD arteries and carotid endarterectomy plaques compared to aged control arteries. The interaction of fibroinflammatory mediators, like CTGF, on atherosclerotic plaque is an important avenue of current discovery. Matricellular proteins, like CTGF, can tune molecular pathways into vicious cycles, and as such, hold promise as therapeutic targets under hostile conditions. Since ECs sense both shear stress and arterial stiffness, it is not surprising that ECs are tuned by this tuning protein. However, the modifiability of plaque biology in the EC-specific CTGF knockout, as well as the CTGF antibody treatment, suggests that manipulation of this pathway is feasible for clinical translation. How this occurs is not yet clear but subject to future investigation. It is important to note limitations of this work that warrant future work. Here we present an animal model of stiffness + d-flow that mimics human PAD. Certainly, no animal model perfectly mimics the human condition. Arterial stiffness and other mechanical parameters differ between mouse and human arteries. However, relative changes between mice and human arteries do appear to be conserved.^47^ Here, we have demonstrated that the Fbln5 KO has stiff arteries that mimic the stiffness of PAD arteries after 4 weeks of d-flow, have baseline endothelial dysfunction, and develop an aggressive atherosclerotic plaque phenotype under atherogenic conditions.

Mechanically, matrix components, collagen and elastin, drive arterial mechanics. Since the Fbln5 KO mimics vascular aging due to a defect in elastic fiber assembling, we were not surprised that unmanipulated KO RCAs mimicked the mechanics of aged arteries. We were excited that the addition of d-flow further stiffened these arteries, and that this stiffening mimicked the ratios seen in PAD arteries compared to their controls. Further, it was surprising to see the similar stair-step upregulation of CTGF in KO compared to WT and d-flow compared to s-flow, and in young/aged and d-flow compared to s-flow arteries, supporting the shared pathways of arterial aging and PAD. The validation of this in d-flow areas of vascular tissue from patients further supports the likely importance of these relationships in PAD. Still, there are additional downstream pathways in our data set that are likely important to the EC contribution to arterial stiffening and flow-mediated atherosclerotic plaque under PAD conditions. It is our hope that others will integrate stiff + d-flow into their discovery science to build a better understanding of therapeutic opportunities in the PAD space.

Matricellular proteins, like CTGF, are of particular interest under PAD conditions, as they may be targeted to dampen signaling pathways important to flow-mediated stiffening and progressive atherosclerotic plaque progression in a favorable manner. Also, relatively little is known about the sex-based differences in EC plasticity, particularly if there is a sex-based difference that is cued by stiffness and d-flow. This work has not exhaustively proven that CTGF is not important to d-flow mediated ASD in females. This is an area of active discovery in our laboratory. Interestingly, the lack of difference in atherosclerotic plaque in ECKO^CTGF^ and ECWT^CTGF^ was not due to differences in absolute stiffness under biaxial arterial testing. It still may relate to the relative response of EC to stiffness + d-flow, particularly as it relates to EC plasticity pathways, but this remains to be proven.

In summary, this work characterized a novel model of PAD mechanics, identified unique molecular signatures related to PAD mechanics that involve precocious EC plasticity, identified an important role for CTGF in d-flow mediated arterial remodeling and in PAD vasculature, and identified that CTGF inhibition therapies may play a favorable role in limiting the impact of PAD on vascular health and flow-mediated atherosclerotic plaque behavior.

## Data Availability

Resource Availability The data that support the findings of this study are available from the corresponding author upon reasonable request.Â Lead Contact Further information and requests for resources and reagents should be directed to and will be fulfilled by the Lead Contact, Luke Brewster. Materials Availability Further requests for resources and reagents should be directed to and will be fulfilled by the Lead Contact, Luke Brewster. This study did not generate new unique reagents. Data availability The RNA files generated from the study (fibulin-5 KO and WT animals) will be available at the NCBI GEO repository on January 1, 2024, as GEO Submission (accession number GSE222583).

https://www.ncbi.nlm.nih.gov/geo/query/acc.cgi?acc=GSE222583

## Notes

### Competing Interest Statement

The authors have declared no competing interest.

### Funding Statement

This work was supported by awards from the National Institutes of Health to Drs. Brewster, Gleason, and Jo (HL143348). The authors' work is supported by funding from the NIH grants HL119798, HL139757, and HL151358 (to H.J.). H.J. was also supported by Wallace H. Coulter Distinguished Faculty Professorship.

### Author Declarations

Emory University approved IRB protocol numbers: 51432 and 70813

